# The public views of and reactions to the COVID-19 pandemic in England- a qualitative study with diverse ethnicities

**DOI:** 10.1101/2022.02.02.22270238

**Authors:** Cliodna AM McNulty, Eirwen Sides, Amy Thomas, Atiya Kamal, Rowshonara B Syeda, Awatif Kaissi, Donna M Lecky, Mahendra G Patel, Ines Campos-Matos, Rashmi Shukla, Colin Stewart Brown, Manish Pareek, Loretta Sollars, Laura B Nellums, Jane Greenway, Leah Ffion Jones

**Affiliations:** Head of Unit to December 2020, (now retired) Primary Care and Interventions Unit, Public Health England; UK Health Security Agency; Primary Care and Interventions Unit, Public Health England; School of Social Sciences, Birmingham City University; University of Bradford; Office for Health Improvement and Disparities and UK Health Security Agency; Public Health England; Head of Healthcare Associated Infections and Antimicrobial Resistance Division, National Infection Service, Public Health England; Department of Respiratory Sciences, University of Leicester, and is supported by a NIHR Development and Skills Enhancement Award; Young People and Families, Office for Health Improvement & Disparities; School of Medicine, Faculty of Medicine & Health Sciences, University of Nottingham; Behavioural Science and Insights Unit, UK Health Security Agency

**Author notes:** **Corresponding author**: Leah Ffion Jones, PhD, Behavioural Science Team Leader, Behavioural Science and Insights Unit, UK Health Security Agency.

**Keywords:** Qualitative, minority ethnic, COVID-19, pandemic, government, public, attitudes, mental health, needs

## Abstract

**Objectives:** To explore public reactions to the COVID-19 pandemic across diverse ethnic groups.

**Design:** Remote qualitative interviews and focus groups in English or Punjabi. Data were transcribed and analysed through inductive thematic analysis.

**Setting:** England and Wales June-October 2020.

**Participants:** 100 participants from 19 diverse ‘self-identified’ ethnic groups.

**Results:** Dismay, frustration and altruism were reported across all ethnic groups during the first six to nine months of the COVID-19 pandemic. Dismay was caused by participants’ reported individual, family and community risks, and loss of support networks. Frustration was caused by reported lack of recognition of the efforts of minority ethnic groups (MEGs), inaction by government to address COVID-19 and inequalities, rule breaking by government advisors, changing government rules around: border controls, personal protective equipment, social distancing, eating out, and perceived poor communication around COVID-19 and the Public Health England (PHE) COVID-19 disparities report (leading to reported increased racism and social isolation). Altruism was felt by all, in the resilience of NHS staff and their communities and families pulling together. Data, participants suggested actions, and the Behaviour Change Wheel informed suggested interventions and policies to help control COVID-19.

**Conclusion:** To maintain public trust, it is imperative that governmental bodies consider vulnerable groups, producing clear COVID-19 control guidance with contingency, fiscal, service provision and communication policies for the next rise in COVID-19 cases. This needs to be combined with public interventions including information, education, modelling and enablement of infection prevention through local community involvement and persuasion techniques or incentivisation. Government policy needs to review and include town and social planning leading to environmental restructuring that facilitates infection prevention control. This includes easy access to hand-washing facilities in homes, work, all food providers and shopping centres; toilet facilities as our Travellers mentioned, and adequate living accommodation and work environment facilitating IPC for all.

**Strengths and limitations:**

- This is amongst the largest qualitative studies on attitudes to the COVID-19 pandemic in the UK general public across ethnic groups, ages and religions, adding insights to previous smaller qualitative studies, from a broader range of participants.
- The qualitative methodology allowed us to discuss participants’ responses around the COVID-19 pandemic, probing their answers to obtain detailed data to inform needs across ethnic groups.
- Most data collection was undertaken in English and therefore excludes non-English speaking sectors of the population who may have experienced the COVID-19 pandemic differently.
- We did not obtain the views of older members of the population over 70 years, who were most at risk.
- The data reflect public perceptions six to nine months into the pandemic when some of the social distancing rules had been relaxed in England; as the pandemic progresses attitudes and needs may well change.

## INTRODUCTION

Death rates from COVID-19 have been higher in the United Kingdom (UK) than many other countries worldwide.^1^ The 2020 Public Health England (PHE) COVID-19 disparities report indicated that the risk of dying from COVID-19 was greater in the over 80s, those living in deprived areas, and 10-200% higher amongst different minority ethnic groups (MEGs) relative to white ethnic groups.^2^ Diabetes was mentioned as a comorbidity on COVID-19 related death certificates twice as commonly in Asian and black compared to white patients.^2^ Surveillance data and systematic reviews confirm the higher mortality in MEGs and areas of deprivation.^3 4 5^ In a PHE rapid literature review and Skype listening events with national, regional and local MEGs, stakeholders expressed deep dismay, anger, loss and fear in their communities about the emerging data and realities of MEGs being harder hit by COVID-19.^6^ In their view, COVID-19 did not create health inequalities, but rather the pandemic exposed and exacerbated longstanding health inequalities affecting MEG communities in the UK.^6^ The 2020 and ‘Build Back Fairer’, COVID-19 Marmot Reviews, indicated that over the last ten years health inequalities in MEGs have grown and health improvements have slowed.^7 8^ The causes of the disproportionate impact of COVID-19 on MEGs are multifaceted and may include: geographical area, living conditions, culture, employment, economic status, and other biological and health related factors; the solutions to it may be just as multifaceted.^9^ The present study aimed to answer the PHE review recommendation for detailed research across a range of ethnic groups and religions to explore in depth their emotions and reactions, attitudes and behaviours in relation to COVID-19,^6^ to understand barriers and facilitators around COVID-19 infection prevention control (IPC), and explore views on what could be done to control the pandemic and reduce health inequalities.

### Methods

This study has been reported in accordance with the Consolidated criteria for reporting qualitative research.^10^ It forms part of a larger study which also explored the attitudes and beliefs of diverse ethnic groups in the UK towards COVID-19 testing and vaccination. [^11^]

#### Topic guide development

The semi-structured interview guide was informed by the 2020 PHE review of disparities in risks and outcomes for COVID-19,^2^ and the Theoretical Domains Framework (TDF) which has fourteen domains that help understand an individual’s behaviour.^12^ The areas covered in the topic guide which mapped to the fourteen TDF domains are shown in Box 1.The topic guide was used flexibly and iteratively with probing of questions.

##### Box 1

**Topics explored in the focus groups and interviews mapped onto the Theoretical Domains Framework**

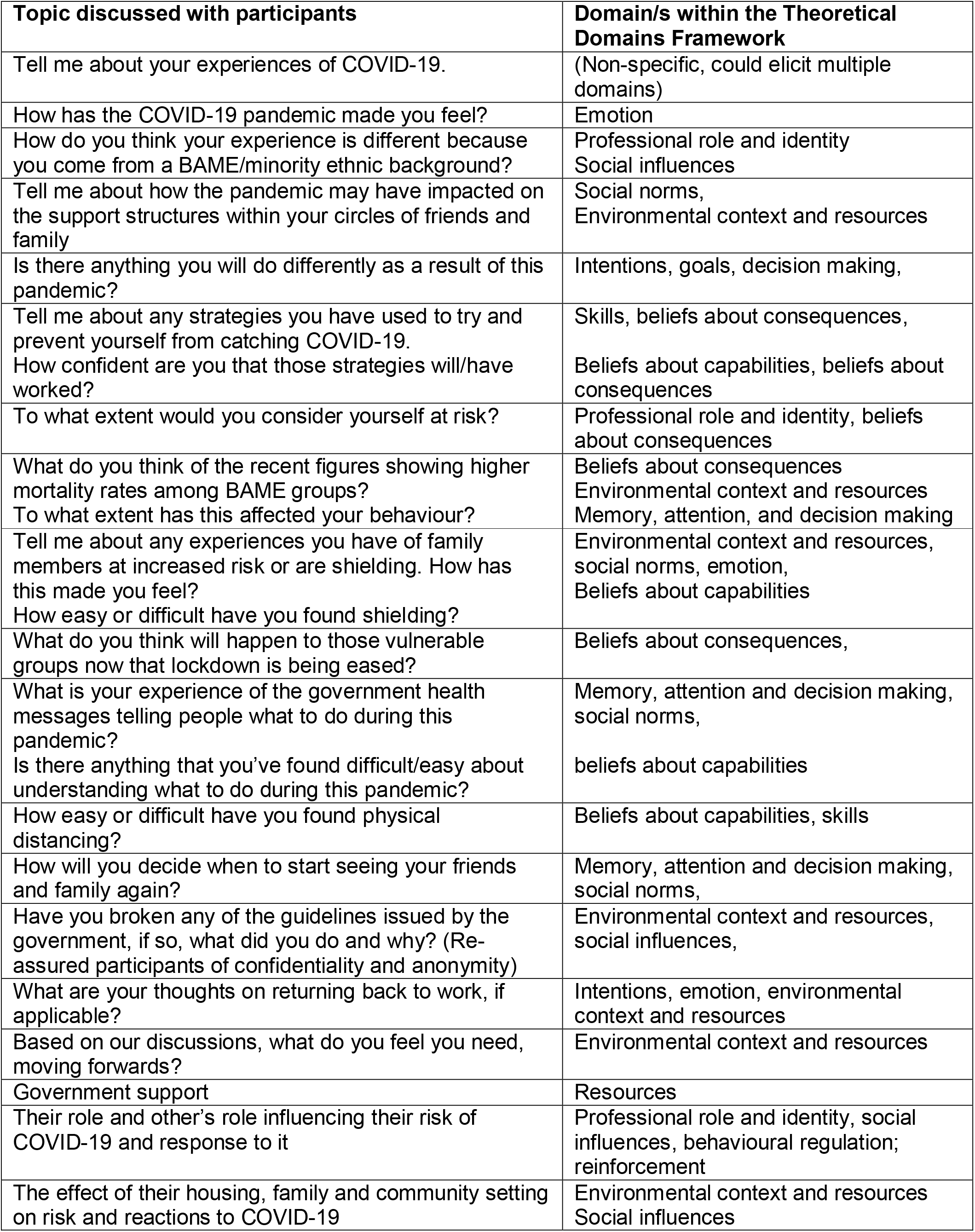

#### Recruitment

One hundred participants were recruited from June to October 2020 (six to nine months into the pandemic) targeting a diverse cross-section of the UK population [^11^] across England and Wales, including diverse MEGs and white British, religions, and occupations. Participants were recruited via the PHE People’s Panel, adverts in Twitter, social media, COVID-19 charities and minority ethnic support groups with chain-referral sampling.^13^ Participants needed a reasonable level of spoken English or Punjabi; they were offered £25 each for their time and contribution.

#### Data Collection

Focus groups (FGs) were conducted via Skype in English, with or without video, with a facilitator and research assistant who took field notes; discussions lasted approximately 60 minutes. Skype interviews were conducted in Punjabi by one researcher. FGs and interviews were audio recorded, transcribed verbatim. Interviews were translated from Punjabi to English; all transcripts were checked for accuracy. Findings were discussed at the end of each FG, and weekly by researchers. Data collection stopped when a range of ethnic and religious groups were recruited, and no new major themes were being identified.

#### Data analysis

Three researchers analysed data inductively using thematic analysis with QSR NVivo.^14^ Twelve transcripts (44%) were double coded and a coding consensus was reached through discussions. Themes were identified from the data, agreed with the steering group, and finalised in a workshop. Overarching themes were created, and with the Michie Behavioural Change Wheel^15^ data was then used to suggest policies and interventions which may help reduce COVID-19 and MEG inequalities. Representative quotes were chosen to expound the themes.

#### Research Group

The research team and steering group included a member of the public, and researchers and healthcare professionals (HCPs) experienced in qualitative research, behavioural science, minority ethnic health, public health including outbreak control and medical microbiology, infectious diseases, and guidance and intervention development.

#### Patient and public involvement

A member of the public was involved in the study steering group from the conception of the study. They also inputted into the design and methodology, as well as data collection tools and recruitment strategy.

### Ethical approval

This study was reviewed and approved by Public Health England’s Research Ethics and Governance Group; NR0215.

## Results

Participants reactions to the pandemic are structured into three over-arching emotional themes: dismay, frustration and altruism displayed across all ethnic groups. These themes along with recommended interventions and policies can be seen in figure 1. Additional participant quotes are provided in a supplementary file.

**Figure 1:**
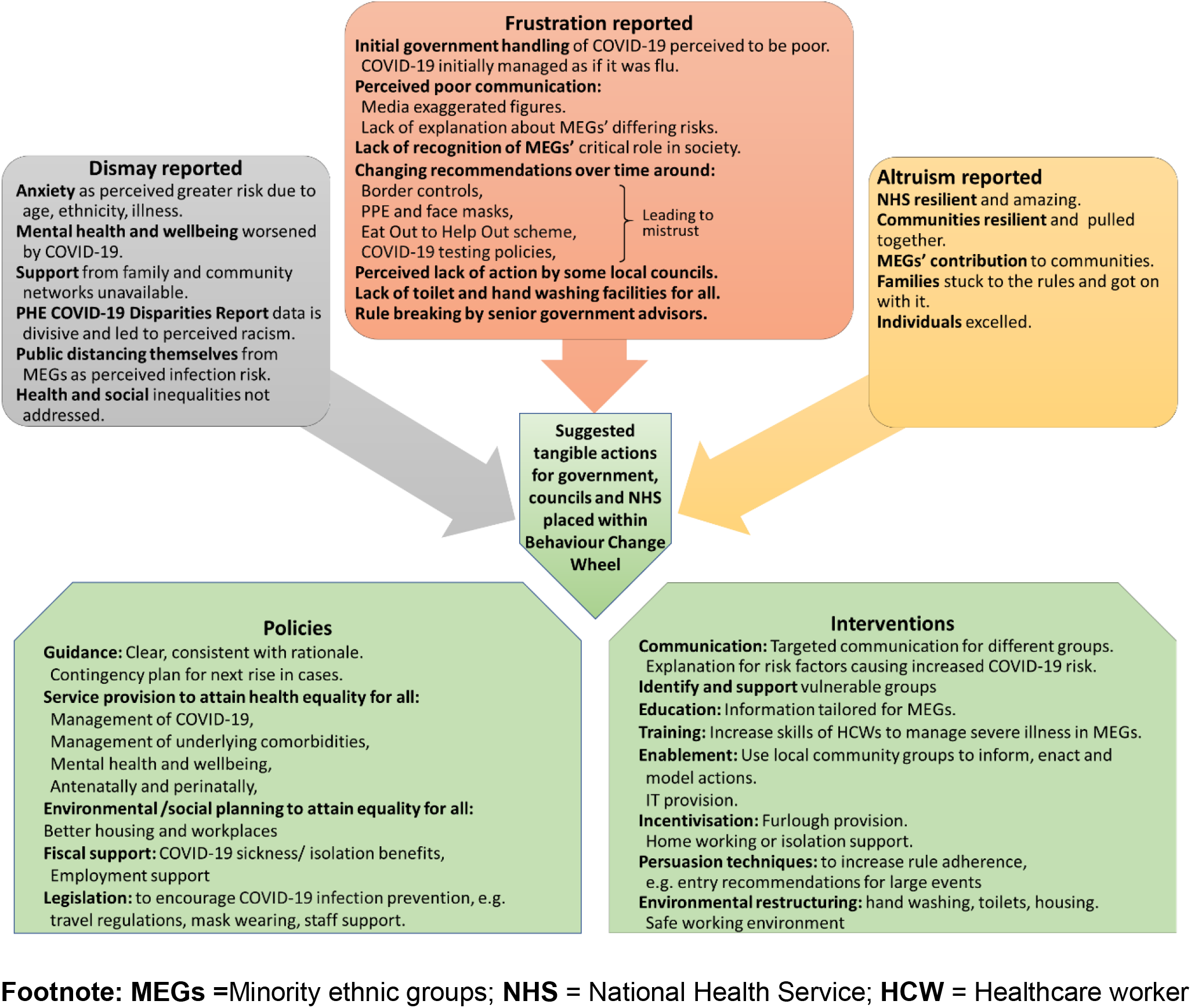
Reported factors contributing to the frustration, dismay and altruism discussed by participants, and suggested actions placed within the behaviour change model^14^ as policies and interventions.

### DISMAY

#### Anxiety due to perceived risk of COVID-19

Many MEG participants mentioned their dismay, anxiety or fear about being at increased risk of COVID-19 and severe illness. Several described themselves at ‘*increased vulnerability’* to COVID-19 so that many had ‘*not left the house since lockdown.’* Risk factors described by all groups included age, and/or blood pressure, diabetes, obesity, and pregnancy. Some participants had avoided their supportive ‘*Community Association meetings and gatherings,’* [FG14, Indian] and others reported that they were ‘*going to be* [very] *cautious until we get a vaccine or until the infection is no longer around*.’ [FG4, SE Asian] Participants highlighted that within a single community there are a range of housing and family units. Those in multiresident or multigenerational households were concerned that they were ‘*exposed to more* [COVID-19] *just because there’s more of you going out and about*.’ [Mixed ethnicity FG1]. This concern about increased risk was echoed by those in public facing roles especially health professionals and public transport.

##### Effect of the pandemic on health and mental wellbeing

Several participants reported dismay at not receiving treatment for other illnesses: ‘*Then I’ve got a lot of, abdominal pain and now the worst one is a chest pain but I cannot get a treatment, they tell me you’re clinically vulnerable, you’re high risk, you cannot come to a medical facility. And there are certain illnesses that you cannot diagnose just over the phone. I was supposed to go for endoscopy, no date* [in September], *and now when you hear news that you may not be allowed to go to A&E, you have to ring 111. And to be honest I’ve heard of two people who have died in their own houses because they cannot access other treatment, that’s a big challenge.’* [FG23 male].

Some MEG participants mentioned that since the pandemic ‘*When I’ve gone to the shops, especially earlier in the pandemic and the lockdown, people would physically step away from myself and I think I’ve noticed that more currently now than it was before’*. [FG7, Chinese]. Several participants reported feeling depressed because of ‘lockdown’ measures. The PHE report highlighting the greater infection rate and mortality from COVID-19 in ethnic minorities, and media response to it, had made MEG participants nervous about any social interaction. ‘*I’ve never experienced anything like this in my life, even though I want to go out, I’m too scared to*’ [FG9, Indian] and this had affected their mental wellbeing, *‘I started feeling sometimes depressed or not productive or just everything’s messed up.’* [FG13, Arab]. MEGs reported the big negative impact that the loss of their support networks had had on their wellbeing *‘I think the Asian community, the Hindu community, the temples and the mosque are a big impact on their family and their friends.’* [FG18, Indian]. ‘*What I have experienced within this period was worse than any time simply because I don’t have family to connect to. I had some workers that I have been seeing face to face. When lockdown came that was not possible, there is no support group meetings or activities that you can go to. So, I felt so isolated in a way that I have never felt to the point it just brought about all the bad memories and including acting on suicidal.’* [FG23 Male]. One participant perceived they had lost their job through mental illness causing a short work absence during the pandemic.

##### Response to increased risk of MEGs contracting and dying of COVID-19

Most participants had heard news showing the greater risk of MEGs contracting and dying of COVID-19; some had specifically heard about the PHE COVID-19 report. MEG participants stated that the report was divisive: ‘*The government have already divided us, why mention ethnic minority?*’ [FG16, black mixed]. This had led to MEGs being kept at a distance by *‘non-ethnic minority people* [as] *they feel more scared coming close to me’* due to fear of infection. [FG15, Bangladeshi]. Jewish participants had become less anxious since they realised that the poorer outcomes in ethnic minorities did not include them. Participants stated that the report increased the perception of risk to some people who had not initially considered themselves at risk of COVID-19 resulting in greater adherence to lockdown rules: ‘*when it came out that BAME* (black and minority ethnic groups) *was worst affected it sharpened people’s radars and there were less meetings and stuff.’* [FG9, Indian].

Some white British participants ‘*felt really a bit saddened’* by the report, stating it made them reflect on *‘the differences, is it kind of genetic? Or is it because people from minority ethnic backgrounds might be more socially deprived or have less access to better health care?’* And in agreement with MEG participants it dismayed them to think there was still such health inequality, *‘we’re in 2020, you’d think that we’re moving towards a more just society’*. Other white British participants wondered if ‘*lack of communication in the right language’* had contributed to increased infections in MEGs, *‘as there’s not enough media information around to explain the situation even for English speaking people, so I dread to think how well it’s been translated for people who don’t have English as their first language’* [FG22, white British]. Other white British participants reflected that the increased risk of infection for ethnic minorities was due to ‘*a large proportion of ethnic minorities that work within the NHS that would have come into contact with people with coronavirus, and lifestyle differences, with big households all living together in some cases.’* [FG20, white British]. The COVID-19 pandemic highlighted pre-existing inequalities at the same time as the Black Lives Matter Movement (BLMM) gained momentum globally. For many participants, the disproportionate impact of COVID-19 on black communities could be understood in line with the concerns of the BLMM.^16^

### FRUSTRATION

#### Belief that figures have been exaggerated

Some participants from across ethnic groups considered that the reports and other COVID-19 MEG figures had been exaggerated or misrepresented by the media as the ‘*stats were just put as big headlines with no explanation,’* [FG16, black Mixed] or stated ‘*it is just fabricated news that Pakistanis get it more and the English don’t. How is it possible?*’ [Interview 1, Pakistani]. This led to mistrust in the statistics: ‘*I don’t know if I should believe those stats.’* [FG10, Bangladeshi]. Participants considered the deaths hadn’t been explained: *‘There was a bit said in the media eventually, but it wasn’t really done the way it should have been done*,– *for example* [increased infections] *because they were NHS frontline or public facing workers’*. [FG19, South East Asian]. While those with awareness *‘that black and Asian people in particular had certain criteria that was making them more at risk*… *and nothing was being done,’* showed *‘frustration and not so much fear for myself, but for the community, for my parents of course*.’ [FG6, Pakistani]

#### Contribution of ethnic minorities unrecognised

Many MEG participants reported feeling frustrated by government as the huge contribution of the MEG communities ‘*didn’t really get recognised’. And who’s on the frontline, who’s dying more? It’s again, it’s the people from the BAME communities*.’ Participants suggested that greater recognition of MEG communities’ contribution, *‘would actually make everyone feel this is such a great time to be united rather than divided*’ [FG9, Indian], and build greater confidence. Some wondered if there was ‘*politics behind this, when they mention the word ethnic minority*,’ [FG15, Bangladeshi] ‘*and what’s been happening elsewhere with the right-wing politics of it all.’* [FG19, South East Asian]

#### Government handling of the COVID-19 pandemic

Participants were mostly negative about the role of government in controlling the pandemic. One participant wondered if the government through their inaction ‘*were deliberately spreading the virus among us* [the general public]. *I think they wanted to like 60% to be affected by Corona so that their immune system becomes stronger and they* [the public] *can, protect themselves.’* [FG15, Bangladeshi]. Another reported *‘the UK government doesn’t seem to give a toss.’* [FG17, Chinese]. Another expressed that government should have managed this pandemic much more seriously and used the experience of ‘*scientists from an Asian background who have experienced SARS in the past.’* [Mixed ethnicity FG3]. Participants mentioned actions the government could have done to slow the pandemic: ‘*why have they started the quarantine of people coming into the country now* [August 2020] *and not three, four months’ ago’*. [FG15, Bangladeshi]. Others reflected that although the government had reacted slowly and made mistakes in the beginning of the pandemic *‘they have now* [in August 2020] *realised that it is really spreading very fast, and they are now taking all possible steps to stop this virus very quickly.’* [FG14, Indian].

#### Belief that messaging is confusing and inconsistent

‘F*rustration is the best word that describes the way it was received by us. In terms of the messages, they were always very, unmatched to the situation. The Prime Minister kept stating like he’s backing up everything with science, but I felt like he was very selective in choosing the messages that he was getting across.’* [FG11, Euorpeans]. Most participants found the government messaging ‘*really confusing,’* and contradictory. ‘*I’m a chairman of a patient’s group and I get a lot of phone calls*, [asking] *what shall I do, etc. There is no clarity*.’ [FG18, Indian]. This was especially as participants perceived that the advice changed so much from *‘You can go to work, - don’t go to work, - stay at home*.’ [Mixed ethnicity FG1]. Inconsistencies caused mistrust; early in the pandemic the ‘*public were told the science doesn’t support us wearing face masks,’* but *‘only recently they’ve been telling us to go out and start wearing face masks, I feel like there’s a bit of a trust issue.’* [FG10, Bangladeshi].

The changing guidance around personal protective equipment (PPE) was a frustration. *‘In my industry, the biggest issue we had was PPE. At one point it was “We don’t need face masks” and then all of a sudden, 12 weeks after the lockdown, “everyone has to wear face masks.”*‘ [Mixed ethnicity FG3, train driver]. The ‘*Eat Out to Help Out’* scheme in England in August 2020 was also particularly highlighted by several participants as being inconsistent with IPC rules in other venues, and likely to increase viral spread: *‘You can see outside restaurants is a long queue of people waiting. So, they’re going to be spreading the virus.’* [FG15, Bangladeshi]. *‘I was in this pub and it was absolutely packed, no masks whatever, and then I walked to the supermarket and it’s masks, wide aisles and there’s like a* [rule for] *how many people can go in, it just doesn’t add up.’* [FG22, white British]. Participants were ‘*really annoyed’* by the government defence of an advisor who broke the rules; stating: ‘*well we’ve literally been following your guidelines to a T and you’ve excused this behaviour, it was a bit of a kick in the teeth.’* [mixed ethnicity FG2].

#### Attitudes towards Public Health England

Participants had some criticisms of PHE indicating *‘that Public Health England had failed to provide timely, accurate information early enough,’* but wondered *‘whether there’s been political pressure at the top to downplay things and to treat this as Influenza, which it isn’t.’* [Mixed ethnicity FG3]. Others considered *‘Public Health England is doing the best they can but politicians being politicians they scapegoat everybody but themselves.’* [FG17, Chinese].

#### Attitudes towards local government

Although there was one positive comment on council support, others reflected that communities did not feel supported by their local councils. The council ‘*is for the local people and they’re not putting their effort in to help the town*.’ [FG18, Indian]. The Traveller community participants were particularly critical of councils stating that ‘*they pass the buck and then nothing gets sorted’*, [FG24, Traveller], reporting they failed to allow access to the public toilet with a wash hand basin, which had been closed initially in the pandemic, even though they were in a ‘*wealthy progressive city.’*

### ALTRUISM

Most participants were very positive about the NHS, stating from their own experiences ‘*I can’t fault the NHS, they’ve done all they can,’* [FG22, white British]; one participant reported negative experiences around the care of her mother with COVID-19. The MEG participants expressed that *‘people from the* [MEG] *communities were proud that their members were on the frontline’* and that individuals and MEGs had made a great difference in their local communities ‘*helping out door to door with services, providing, especially where the elderly couldn’t go’*. Most participants from all groups followed the lockdown and social distancing rules stating ‘*when a situation like this arises, that’s what you have to do. If you’re being told you have to do this then so be it, you just have to go with it.’* [Mixed ethnicity FG5] Others commented how the pandemic had ‘*brought out the good in people and how they could work as a community’*. [FG13, Arab].

### TANGIBLE ACTIONS REQUESTED BY PARTICIPANTS

Participants across all ethnic groups wanted to see similar ‘*tangible actions*’ to prevent the pandemic worsening. Using the Behaviour Change Wheel these suggested actions were placed within government policy changes and interventions that would help control the pandemic and communities.

### POLICIES

#### Policies that increase social equity

Most groups wanted to see the government taking long-term action to address inequalities in terms of class, ethnicity and race, because *‘until you’ve got equity and equality in terms of class and racism, I think you’re always going to have health inequality.’* [FG6, Pakistani].

#### Healthcare policies to increase equity across ethnic groups

Participants indicated that the term BAME tended to group all ethnicities together, but their needs, co-morbidities and risk of COVID-19 and death were very different. ‘*The NHS clearly needs to change, and it needs to be much more specific with how it deals with different races. Black people are different from Asian people, or different from Chinese people, and this BAME thing doesn’t work*.’ [Mixed ethnicity FG3]. For example, ‘*there’s a 20% increase in diabetes within the South Asian community, so we need to tackle tha*t.’ [FG18, Indian]. Other major disparities in health morbidity were highlighted including maternal mortality. *‘I always kind of knew about the stats that black women are five times more likely to die in childbirth than white women.’* Perinatal risks were considered greater during the pandemic as partners were only allowed to support mothers in the final stages of labour. *‘I think you need a partner there to advocate for you* [throughout] *and support you. So, I do really feel sorry for women giving birth without their partner.’* [Mixed ethnicity FG2].

#### Guidance

All participants requested clear guidance to reduce confusion: ‘*we need some clear guidance, and more clarity from the government, in a way that people can digest.’* Participants also requested a rationale for any guidance or policies as ‘*transparency* [from government] *moving forward, means that people can make more informed decisions about how to behave and protect themselves and their family’* [Black ethnicities FG8]. Alongside this, participants wanted ‘*accountability with things that went wrong*.’ [mixed ethnicity FG2].

Participants stressed that one set of guidance or information source does not fit all ethnic groups or all the individuals within an ethnic group. ‘*On one end of the spectrum you may have ethnic minorities in low socioeconomic status groups, but on the other end you’ll have people who are very well educated, who are the key workers, who are the doctors and nurses and a lot of ethnic minority people fall into that category as well.’* [Mixed ethnicity FG5]

A Jewish participant approved of the fact that their religion was specifically recognised in COVID-19 guidance for a Jewish Festival, noting *‘I think somebody has actually given it some thought as to how we function as a religion, and I find that quite comforting.’* [FG21, Jewish]. Although other Jewish participants stated that they didn’t need *‘specific support from the government just because I’m Jewish, because otherwise they’re going to have to single out every other religion as well. I’d say that I would probably just do the same as every religion,’* [FG21, Jewish], indicating that guidance should be relevant to all, religious or not.

#### Support of employment

Participants indicated that ‘*when the Government launched the furlough scheme, our company was really happy and they wanted to keep everyone to make sure everyone is working, nobody is unemployed*. [FG15, Bangladeshi]. However unemployment issues were *‘going to be hard over such a long time*’ and that all ethnic groups ‘*would need support to get back to work*, …*via a recruitment office or anything like that*.’ [FG13, Arab]

### INTERVENTIONS

#### Communication through different channels

participants requested *‘more campaigns educating the public, to disseminate information that is readable and digestible, because a lot of the time most people just get their information from the media. Tackling that would really help people build confidence in the government and the organisations that are responsible for looking after their health.’* [Black ethnicities FG8]. One participant highlighted the importance of local ethnic minority radio stations and community leaders to get across appropriate messaging. Another highlighted the need for IT support for lower income groups as *‘they don’t have access to technology, some don’t know how to use technology, some of them they don’t have money to have internet in their house. They don’t have a proper equipment to* [join] *their support group now we are online.’* [FG23 Female].

#### Work with local communities to increase understanding and optimise support

MEGs were particularly vocal that to help unite communities they wanted their community groups, local charities and community centres recognised and involved in the development of any policy and intervention forward planning. Participants suggested that: ‘*Outreach at council level would be really helpful for the community right now’* [FG6, Pakistani]. They stressed the importance of utilising the learning from local community leaders about ‘*the grass roots people that you need to connect with’*, and to have *‘a contingency plan ready and waiting if the second wave does come.’* [FG19, South East Asian].

#### Environmental restructuring

Traveller participants reported that during the COVID-19 pandemic the government were not providing for their basic needs: ‘*it should be automatic that every authority, should go out and provide a water bowser toilet, access to gas and wood’*. [FG24, Traveller] Others highlighted that multiresident households and public-facing workers needed to be considered in policies. *‘But the real question is, how can we avoid the communities of getting infected more. If I visit them in houses, I’ve seen flats where there are two different families with two or three kids each sharing three rooms. I don’t see any sides ready to do what it really takes. The answer is systematic, … in order to actually do something really, really productive for them.’* [FG11 male]

##### Training for Healthcare workers around managing critical illness in different ethnic groups

Participants worried that *‘there is a big difference in the way healthcare workers look at patients who are BAME, ‘we’re not taught how things manifest differently because of the colour of your skin, so I think that’s why a lot of the time people are mis*[treated]. *And I think that’s a big thing with COVID as well*,..*we’re not taught the differences so, we have to go out of our way to see the differences, so I think that* [training] *would be a really good thing that Public Health could make sure happens.’* [Mixed ethnicity FG2, medical student]

#### Interventions to increase adherence to rules

Many participants across ethnicities discussed the need for greater ‘*enforcement and mandating* [of lockdown] *rules*. The interviews had taken place shortly after a key government advisor had broken COVID-19 rules and there had been no repercussions for them. Many participants across ethnicities considered that there should be *‘heavier fines or something for those who are breaking the COVID rules’*, [FG14, Indian] with something *‘in place so people actually believe that something will happen to them if they don’t abide.’* [FG20, white British]. Many reported that younger individuals were following the COVID-19 guidance less rigorously than others at risk of severe infection. *‘Policing the situation hasn’t been right from the beginning, they* [the police*] could have been stricter everywhere, stricter in the inner-city area where* [young] *people are gathering together as they’re behaving like there’s nothing happening.’* [ FG14, Indian]. They discussed that there needed to be equality of policing across all levels including those in senior government roles.

## Discussion

### Key findings

Dismay, frustration, and altruism were reported by all our participants during the first six to nine months of the COVID-19 pandemic. They considered that government handling of and communication around the pandemic had been poor. The PHE COVID-19 disparities report, had exacerbated this dismay and frustration as the disparities in mortality had not been explained well, had amplified the insufficient action to reduce health inequalities, and these perceptions just ostracised MEGs more. Participants indicated that government policies should include clear COVID-19 guidance, with a contingency plan for the next rise in cases; information and education tailored for local communities, MEGs, and health care workers (to improve management of severely ill MEG patients); clear communication and social marketing, and enablement of the population through supporting local communities to help inform and enact interventions.

#### Comparison with existing literature

##### Social stigma and discrimination

The social stigma and discrimination reported by many MEG participants in this study is concerning, and aligns with reports from health workers, people coming from abroad, and those in quarantine.^17^ Community leaders in Canada identified that poor working conditions facilitated viral spread among marginalised groups, and the role of anti-black racism in response to higher rates in MEGs.^18^ Our participants also noted the importance of policies that reflect the lived experiences of racialised populations in a meaningful way. The Canadian study also referred to the BLMM and suggested it was an opportunity to change towards equity and fairness in health, and overall quality of life.^18^ In June 2020, the City of Toronto declared anti-black racism a public health crisis, reprioritising city resources to address this issue during COVID-19 recovery planning.^19^ Our participants however were less positive about future likely changes, judging the government by its perceived current lack of action towards equality. In June 2020 the Minister for Equalities was tasked with addressing specific issues in response to the publication of the Disparities review.^20^

##### Health

Many of our participants discussed health inequalities in the UK and stated this had to be a priority to address. Our participants’ request for more information for groups with comorbidities to help explain their increased risks and what they can do, has been echoed in other studies, including carers of diabetic children and pregnant women.^21 17 22 23^ General practitioners across Europe have expressed their concern about ‘collateral damage’ resulting from routine care being postponed or limited due to COVID-19;^24 25^ this effect is likely to be greater in ethnic minorities who already have poorer healthcare access. The social isolation and closure of services exacerbated social and mental health problems across all ethnic groups in our study and others,^26 23^ but especially effects marginalised and vulnerable groups.^27 28^ Mental health and wellbeing are a common motive for breaking social distancing guidance in the UK.^29^ A 2021 UK review suggests that changes in overall population mental health since the beginning of the pandemic were greater for ‘non-whites,’ younger adults (aged 18 to 30), and women.^30^ The <u>UK Biobank</u> indicates that people with a psychiatric disorder have greater COVID-19 mortality. It is important to identify the vulnerable, and groups at greater risk of mental illness and ensure support through the pandemic.

Several participants in our study were worried about the increased risk of perinatal death with women experiencing labour unsupported during COVID-19. Maternal deaths in the UK are still two times higher in Asian and four times higher in black than white women;^31^ clinicians should have a lower threshold to review, admit and consider multidisciplinary escalation of symptoms in women of MEG background, this is particularly true during the pandemic.

##### Guidance

Our participants discussed the need for clear transparent guidance, as they like others, were frustrated by changing rules.^29 32^ Constantly changing news and information in Australia led to public confusion and distress.^33^ Additionally, research indicates that changing guidance, breaches of lockdown among influential figures or communicating with unwarranted certainty around COVID-19 or vaccination leads to less compliance and/or trust in the information by readers compared to providing consistent information, and including uncertainties.^34 29 35 36^

##### Environmental and social policy

Participants discussed how a multiparticipant/multigenerational household and/or work environment and social interactions put them or others at increased risk of acquiring COVID-19. In a March 2020 UK survey, MEG participants and others from more disadvantaged backgrounds reported they would be less able to work from home or self-isolate if needed, suggesting the existence of structural or financial barriers to adopting preventive behaviours in these groups.^37^ Decreasing the risk of contracting COVID-19 going forward may need a government policy examining town and social planning leading to environmental restructuring facilitating IPC. This includes easy access to hand-washing facilities in homes, work, all food providers and shopping centres; toilet facilities as our Travellers mentioned, and adequate living accommodation and work environment for all.

##### Financial and employment support

The financial pressure of COVID-19 on populations and especially disadvantaged groups, and need to undertake an equality impact assessment of the financial barriers experienced by them, has also been highlighted by others.^38 32 23 17^ By Autumn 2020 when this study was completed the job furlough scheme introduced in March 2020 helped protect *‘at risk jobs across the UK by providing employees with incentives to keep staff employed’*; national data indicated more than half of those furloughed had returned to work by mid-August 2021.^39^ Our participants indicated the likely need for ongoing employment support after the furlough scheme ends to retain jobs.

##### Increasing compliance with COVID-19 rules

Many of our participants expressed a belief that the rules and laws introduced by government should be enforced. However an enforcement-based approach can dilute the publics’ voluntary commitment to comply, so called ‘control aversion’.^40^ German population enforcement lowered likely compliance with COVID-19 social distancing, use of a contact tracing app, and vaccination measures.^40^ Control aversion is greater in younger adults; if there is lack of clear rationale for the rules, or a perception that the government does not trust the general public. ^40^ Thus, although legislation and policing may be appropriate in some instances, it is better avoided and replaced with enablement and support, or incentives.

##### Involve community groups

It is important for communities, government, guidance, advice and the media to harness the achievements of individuals, families, the local community, and the NHS that have been highlighted by our participants and those in other studies.^41^ A qualitative study with COVID-19 control decision makers in Kerala, India, where mortality was lower than other areas of India, showed the backbone of their strategy was community participation and local leadership which mobilised community self-help groups.^42^ Our participants and others stressed involving local community groups in information development to allow different forms of intervention adapted for different population and risk groups^29^ with different opportunities, privileges, access to health care and IPC adherence.^18^ In October 2020 the UK government announced an additional £4m for COVID-19 targeted messaging for MEGs and a new ‘*Community Champions’* scheme to fund work with grassroots advocates, and voluntary and community groups from impacted communities.^43^ The COVID-19 Scientific Pandemic Insights Group on Behaviour (SPI-B) advised where community trust is low community champions can be a key pillar to support IPC measures,^44^ as they can reach and support isolated or marginalised individuals, help communicate health messages,^38 44^ reducing health inequalities. Community champions should be embedded in effective community engagement strategies.^45^

#### Unanswered questions, future research

It is important to obtain the views of those underrepresented in this sample, that is non-English speaking MEGs and over 70 year olds or their carers, as these groups had the greatest morbidity and mortality associated with the pandemic. Service providers, government, PHE successor bodies and councils should be shown the results to obtain their response to the public’ attitudes to handling of the pandemic and explore how they can improve management of the pandemic through its next stage. As the pandemic progresses, further local community exploration with MEGs will be needed to understand the specifics of local situations to optimise policies and interventions.

## Implications

To improve trust and compliance future reports or guidance should clearly explain any stated differences in health outcomes by ethnicity or other risk group, including specific messages for these groups and concrete actions to minimise any risks. Messaging should reflect the uncertainty in data or advice and how guidance may change going forward as new evidence becomes available. A contingency plan is needed to mitigate the impact of COVID-19 across all communities including MEGs, the vulnerable and socially disadvantaged individuals, in preparation for any rise in cases and for future pandemics. To minimise inequalities and racism these plans should promote collective identity and share positive, supportive and non-judgmental communication.^46^ Equality across ethnicities for COVID-19 and healthcare is essential, and the NHS and local communities will need to be supported to attain this. It is imperative that governmental bodies and councils produce clear, consistent, and transparent health, social, fiscal and environmental policies to reduce social and health disparities, informed and actioned by different ethnic and demographic groups nationally and locally.

## Supporting information

Supplementary box of quotes

## Data Availability

Data are available on request.

## Acknowledgements

We would like to extend our thanks to all public representatives, health care professionals, researchers and expert advisors who contributed to this study. Thank you to all participants for providing their time and sharing their views and experiences for FGs and interviews.

## Contributors

- CAMM: had substantial contributions to the design of the work (helped develop protocol, interview questions); reviewed analysis, drafted all versions of the manuscript, critically revising drafts; gave final approval of the version to be published; and has agreed to be accountable for all aspects of the work.
- ES: collected data; had substantial contributions to the analysis and interpretation of the qualitative data; contributed to all drafts of the manuscript providing quotes and interpretation and critically revised it; gave final approval of the version to be published; and has agreed to be accountable for all aspects of the work.
- AT: had substantial contributions to the analysis and interpretation of the qualitative data; critically commented on versions of the manuscript; gave final approval of the version to be published; and has agreed to be accountable for all aspects of the work.
- ATK: had substantial contributions to the design of the study (recruited participants); collected data; had substantial contributions to the analysis and interpretation of the qualitative data; critically commented on versions of the manuscript; gave final approval of the version to be published; and has agreed to be accountable for all aspects of the work.
- RBS: had substantial contributions to the design of the study (development of protocol, drafted interview questions); collected data; critically commented on versions of the manuscript; gave final approval of the version to be published; and has agreed to be accountable for all aspects of the work.
- AK: quality checked data; had contributions to the interpretation of the qualitative data; critically commented on versions of the manuscript; gave final approval of the version to be published; and has agreed to be accountable for all aspects of the work.
- DML: had substantial contributions to the design of the work (helped develop protocol and interview questions); reviewed analysis; critically commented on versions of the manuscript; gave final approval of the version to be published; and has agreed to be accountable for all aspects of the work.
- MGP: had substantial contributions to the design of the study (recruited participants); reviewed analysis; critically commented on versions of the manuscript; gave final approval of the version to be published; and has agreed to be accountable for all aspects of the work.
- ICM: had substantial contributions to the design of the work (reviewed interview questions); reviewed analysis; critically commented on versions of the manuscript; gave final approval of the version to be published; and has agreed to be accountable for all aspects of the work.
- RS: had substantial contributions to the design of the work (reviewed interview questions); reviewed analysis; critically commented on versions of the manuscript; gave final approval of the version to be published; and has agreed to be accountable for all aspects of the work.
- CSB: had substantial contributions to the design of the work (reviewed interview questions); reviewed analysis; critically commented on versions of the manuscript; gave final approval of the version to be published; and has agreed to be accountable for all aspects of the work.
- MP: contributed to the design of the work (reviewed interview questions); reviewed analysis; critically commented on versions of the manuscript; gave final approval of the version to be published; and has agreed to be accountable for all aspects of the work.
- LS: contributed to the design of the work (reviewed interview questions); reviewed analysis; critically commented on versions of the manuscript; gave final approval of the version to be published; and has agreed to be accountable for all aspects of the work.
- LBN: had substantial contributions to the design of the work (reviewed interview questions); reviewed analysis; critically commented on versions of the manuscript; gave final approval of the version to be published; and has agreed to be accountable for all aspects of the work.
- JG: had substantial contributions to the design of the work (reviewed interview questions); reviewed analysis; critically commented on versions of the manuscript; gave final approval of the version to be published; and has agreed to be accountable for all aspects of the work.
- LFJ: project managed study from start to close; led the analysis and interpretation of the qualitative data; had substantial contributions to the design of the study (led on development of protocol, gained ethics approval, drafted interview questions, recruited participants); led the collection of data; had substantial contributions to the analysis and interpretation of the qualitative data; critically commented on versions of the manuscript; gave final approval of the version to be published; and has agreed to be accountable for all aspects of the work.

## Data sharing statement

Data are available on request.

