## Supplementary box of quotes for "The public views of and reactions to the COVID-19 pandemic in England- a qualitative study with diverse ethnicities"

**Supplementary Box: Participant quotes to support the key findings**

| **DISMAY** |
|---|
| **Anxiety due to greater risk of infection**<br>*'So, it's an anxiety that I have maybe put upon myself, but because I know that the levels amongst the South Asian population are highly. And also because I'm (in my 60s), so that's another added concern for me…*' [Female, FG4 June 2020, South East Asian]<br>**Age:** *'…I have got blood pressure, so I'm in a very vulnerable age, so it's just impossible to go out, we have (inaudible) Indian Community Association meetings and gatherings and all that, we can't do any of this thing at the moment…'* [FG14 August 2020, male, Indian]<br>**Other comorbidities increasing risk:** *'…Yeah, I feel at an increased vulnerability I suppose, because of the combination of having recently given birth and my ethnicity as well. [Mixed ethnicity FG2 June 2020, Female] I'm also quite I suppose frustrated with the government, the lack of government response to try to get to the bottom of why it does seem to be affecting BAME communities so much…'* [*Mixed ethnicity FG2 June 2020,* Female]<br>*'…the pandemic has been quite terrifying as far as I'm concerned, because I'm also a diabetic, so slightly overweight, so I've got all the things that make me pretty vulnerable, so I've not left the house really since the lockdown…'* [FG20 September 2020, male, white British]<br>**New MEG mother:** *'I feel at an increased vulnerability I suppose, because of the combination of having recently given birth and my ethnicity as well'* [*Mixed ethnicity FG2, female, June 2020*]<br>**Housing:** *'…especially when you live with eight people, I think you're more, you're exposed to more just because there's more of you going out and about.'* [Mixed ethnicity FG1, June 2020, male]<br>'You've got *different living conditions, for some people who are living with extended families it might be much harder for them to self-isolate'* [FG19 September 2020, South East Asian].<br>**Diet:** *'…It's well known that Asian people eat a lot of packaged food and dried food. And they don't normally do exercise as much as I would say other people. This is generalising.'* [FG18 August 2020, male, Indian]<br>**Public facing roles:** *'…we got a problem of us getting this type of disease. … I put some plastic,* [in] *the middle of the driver to passenger. I spent nearly £150 and in the end it's not worth it. It's quite expensive but I had to do because of the safety. The problem is some customers, most of the customers, they don't use masks sometimes, they're coughing even they don't bother to open window, it's very bad experience and it's not good financially.'* [FG15 August 2020, male, Bangladeshi taxi driver]<br>**Ethnicity:** *'I've isolated myself deliberately and that's because I'm South Asian and because of all the stuff in the news…And again, that psychologically I put it down to the fact that it's because I'm South Asian. So, I'm going to be as cautious as I am being for as long as I possibly can until we get a vaccine or until the infection is no longer around.'* [Female, FG4 June 2020, South East Asian]<br>**Effect of the pandemic on other health issues:**<br>*'Then I've got a lot of, abdominal pain and now the worst one is a chest pain but I cannot get a treatment, I, they tell me you're clinically vulnerable, you're high risk, you cannot come to a medical facility. And the whole thing is over the phone and there are certain illnesses that you cannot diagnose just over the phone. And it's from March that I started out with this pain, up to now I was supposed to go for endoscopy, no date, and it's now* [in September] *focus on COVID and now when you hear news that you may not be allowed to go to A&E, you have to ring 111. And to be honest I've heard of two people who have died in their own houses because they cannot access that treatment, other treatment had been put on hold, that's a big challenge.'* [FG23 September 2020, black African male].<br>**Effect of the pandemic on mental health and wellbeing:**<br>'*I think the Asian community, even the Hindu community, the temples and the mosque are a big impact on their family and their friends, and the meeting, and the welfare…'* [FG18 August 2020, female, Indian]<br>*'I started feeling sometimes depressed or not productive or just everything's messed up or accumulate my tasks'* [FG13 August 2020, female, Arab]. |

| |
|---|
| *'They said they support mental health, there's so many policies and things and OK we'll support* [you]. *But during the pandemic I once had two very hard days that I did not work. I mentioned that, I called* [them at work], *everything, but it was the second time happening during my employment. I noticed every time this happened, although they say they support it but they start a meeting after it mentioning issues about my work which happens only directly after me not working during the days of my hard depression.'*<br>'*What I have experienced within this period was worse than any time simply because I don't have family to connect to. I had some workers that I have been seeing face to face. When lockdown came that was not possible, there is no support group meetings or activities that you can go to. So, I felt so isolated in a way that I have never felt to the point it just brought about all the bad memories and including acting on suicidal.'* [FG23 September 2020, Male Gambian].<br><br>**MEGs feel they are being avoided:** *'When I've gone to the shops, especially earlier in the pandemic and the lockdown, people would physically step away from myself and I think I've noticed that more currently now than it was before.'* [FG7 July 2020, Chinese male].<br>*'So, I stayed in a lot and I don't think I was mentally as happy, because it was quite a big change from being in the lab and being very active, to just staying at home and not doing much, and I would get a lot of anxiety just going for a run because you don't know who you'll encounter, particularly being from an Asian background.'* [FG7 July 2020, female Chinese]. |
| **Response to the PHE COVID-19 review of disparities in risks and outcomes report**:<br>'*I don't consider* [myself] *as a risk but yes when it comes to meeting other people where there is non-ethnic minority, they feel more scared coming close to me. That's what I, because the Government already divided us, by mentioning ethnic minority, that was, they shouldn't have used this word ethnic minority at all you know, in my view.*' [Black ethnicities FG16 August 2020, male].<br>*'I didn't really think much of it. I don't think I was either concerned or stressed or, it, I don't think it affected me in any way. I just accepted that as a report rather than anything else.'* [FG11 July 2020, European female].<br><br>**Jewish participants initially concerned that they were included in the MEGs at risk:** *'…initial stages when there were stories about maybe the Jewish population as being one of the populations that was susceptible to the virus, after that, it was, it just concentrated on the BAME communities. And then it started to feel as if, oh it's all right, it's not me, sort of in that respect…'* [FG21 September 2020, male, Jewish]<br>**Impacts of report on adherence to rules:***: 'I feel like to start off people weren't taking COVID that seriously here, but then when it came out that BAME was worst affected it kind of sharpened people's radars and I think there were less meetings and stuff that shouldn't have been happening*' [FG9 July 2020, Indian, male]<br>**Emotional impact of report and statistics:** *'… I'm afraid that I read in the paper that the BAME community is more vulnerable to coronavirus, so that's, it's a scary time, you don't want really, because of my health, I don't really want to go and meet anybody or, I've never experienced anything, and I'm sure everybody is the same, I've never experienced anything like that in my life, and even though I want to go out, but I'm too scared to go out, because I'm not worried about just myself, I don't want to pass it, if anything does happen I don't want to pass onto my wife or my kids or grandkids, it's, I don't know, I just, scary times…'* [FG9 July 2020, male, Indian]<br>**White British and Jewish participants reflected on the reasons ethnic minorities were more at risk** Saddened : *'…I felt really a bit saddened, by it. It made me think about, what is the social, what is the difference, what are the differences you know, is it kind of the genetic, or is it because people from minority ethnic backgrounds might be kind of more socially deprived or kind of have less access to better health care or you know what is it, and I suppose it makes, it made me think about, we're in 2020, you'd think that we're moving towards kind of a more justified society but because of these set of differences, you know what's behind them kind of thing…'* [FG20 September 2020, male, white British] |

| |
|---|
| *Genetics '…I still think there is some genetic factor in it, I don't know what it is exactly it, …'* [FG21 September 2020, male, Jewish]<br>***Lack of explanation for differences in risks for MEGs***<br>*'…I just wondered if it was sort of a lack of communication in the right language because where I'm from there's, like South East London, there's quite a high proportion of ethnic minority communities where I live and I mean, I've been in doctors surgeries where they have to bring their children in with them just to like translate so, I think if there's not enough media around to explain the situation even for English speaking people, then I sort of, yeah, dread to think how well it's been translated to people that can't actually, who don't have English as their first language…'* [FG22 September 2020, white British]<br>*Increased risk through work or lifestyle'…We need to consider that there are a large proportion of ethnic minorities that work within the NHS that would have come into contact with people with coronavirus. There are lifestyle differences, big households all living together in some cases.* [FG20 September 2020, white British]<br><br>**The black lives matter movement**<br>*'So there were all these issues were what people in the community talk about as well, especially the ones that are a bit more intellectual, they're the kind of conversations why, why are we still, why are the ethnic communities still treated third rate? And you've only got to look at what's happened with the Black Lives Matter, all this what's going on. And it just, the thinking, the mentality of people that's thinking, are we really going to get there?'* [FG19 September 2020, South East Asian]<br>*'…I don't know whether it's all in line with the Black Lives Matter message, but people of ethnic minorities are living in more deprived conditions and then it's almost like using it as a bit of a blame tool for, the country's not being fair or inclusive of these other minorities and that's why they're all dying, because they're not given the opportunities that white people are…'* [FG20 September 2020, white British]<br>*'…the recent Black Lives Matter movement, I would have liked to have gotten involved in that but felt that I couldn't really because of the social distancing, so I would have liked to have attended a protest, we had a protest but couldn't and that did leave me with feeling a bit, almost I do feel quite guilty about that I suppose, but quite conflicted at the same time…'* [*Mixed ethnicity FG2 June 2020, female*]. |
| **FRUSTRATION** |
| **Belief that figures have been exaggerated:**<br>'…*So what annoyed me most was the stats were just put as big headlines with no explanation, no reporting or even just say, we don't know why this happened, we're going to investigate. But I just thought it was just another case of the press and everybody else just putting stuff out there to, basically to make news. And that annoyed me because what I also thought was that would lead to prejudice and people avoiding black and ethnic minorities, and all this sort of stuff, because, again, it's disinformation, is the way I saw it* . [FG16 August 2020, black Mixed]<br>**Uncertainty that statistics were true:***'What proof do they have that the Asians get it more and the English don't? How is it possible? The germ can attack anybody. Why should it be more in Pakistanis? ... It is just fabricated news that Pakistanis get it more and the English don't. How is it possible?'* [Interview 1 October 2020, female, Pakistani].<br>*'A cynic of statistics, I really am. Because people can make them seem and do whatever they want.'* [FG20 September 2020, male, white British]<br>**Data wasn't explained, leading to confusion:** *'There were a bit said in the media eventually, but that's that, so they were never really done, or it was done, but it wasn't really done the way it should have been done.'* [FG19 September 2020, South East Asian]<br>*'…I don't know what stats to believe because I think someone on the call was saying about the news headlines, about the Bangladeshi community, because I come from a Bangladeshi community so there's a high link for people in BAME community, Bangladeshi community to have COVID. So, I don't know if I should believe those stats or, yes, I don't know. So, I'm just a bit confused I think, it's just mixed emotions and feelings and experiences.'* [FG10 July 2020, female, Bangladeshi]<br>**No action following reports of increased risk to MEGs:***'I think just having that awareness of the scientific literature that was coming up, quite early on there was, from, I'd say months' ago, there was an awareness of black and Asian people in particular* |

*having certain criteria that was making them more at risk. And again, that comes, there was various comorbidities but, age, diabetes, etc, and there was an awareness of that, and nothing was being done. So, there was, yes, a lot of frustration and not so much fear for myself, but for the community, for my parents of course, so yes, it was frustrating definitely and a bit of fear I guess, yes.'* [FG6 June 2020, female, Pakistani]

*I'm also quite I suppose frustrated with the government, the lack of government response to try to get to the bottom of why it does seem to be affecting BAME communities so much…'* [*Mixed ethnicity FG2 June 2020,* Female]

**Government have not done enough to address longstanding inequalities in MEGs:** *'I think a lot of the experiences of BAME people are very unheard and haven't really been looked at by the government. I think health inequality is obviously something that's existed for a long time. I think the government knew probably, or should have known, realistically, that this would have an effect on BAME people. They didn't do anything about. The report came out, they still didn't do anything about it…'* [FG9 July 2020,female, Indian]

*'As you've mentioned, the BAME report came out. They've said this, that, that, but they haven't given us what needs to be done. To me, that is a very important factor. What other, what measures are they going to put in place?* [FG18 August 2020, male, Indian]

*'…So, I sometimes feel there is a politics behind this when they mention the word ethnic minority.'* [FG15 August 2020, male, Bangladeshi]

*'…when there was a lot of racism what's been going on, with the government viewed as more to the right, and what's been happening elsewhere with the right-wing politics of it all.* [FG19 September 2020, South East Asian]…

**Contribution of ethnic minorities unrecognised**:

*'And people from the communities were … were annoyed, they were angered as well, why? We're here, we're part of the society, we're part of this community, we are British, we are part of this country, we get all this negativity. And who's on the frontline, who's dying more? It's again, it's the people from the BAME communities, which didn't really get recognised.'* [FG19 September 2020, South East Asian]…

*'Helping out door to door with services, providing items where, especially the elderly couldn't go, so I felt like this should have been covered more by the media as well and the government as well should have recognised these things, that would actually make everyone feel like, OK, you know what, the, this is such a great time to be united rather than divide, be divided'…*[FG9 July 2020, Indian],

*' it's only now in the last month or so, that we've got some recognition from the local council. We did a lot of stuff off our own backs, it really was a key element in getting messages out. So, because there was not much taxying, I was volunteering. … We had another project where we were giving cooked food out to all the people who couldn't access food. -they were all communities, it wasn't just like BAME or specific communities.'* [FG19 September 2020, South East Asian]

**Government handling of the COVID-19 pandemic**:

*'…I think the Government deliberately spread this virus among us.*, *I think they wanted to like 60% to be affected by Corona so that their immune system becomes stronger and they* [the public] *can protect themselves.*

*Now, number one, why did they stop, or started the quarantine of people coming into the country now and not three, four months' ago, when they realised, hang on, I mean I'm sure we could have done something like that, that's number one.'* [FG15 August 2020, male Bangladeshi]

*'Only the UK government doesn't seem to give a toss.'* [FG17 August 2020, Chinese]

*'In the beginning, the government did make some errors in a way that they did not close the border when this virus started spreading, and there was no check at the Heathrow Airport, even now there is hardly any check at the Heathrow Airport, people just walk in from any of the country, …'* [FG14 August 2020, Indian]

**Positive reflection about government:***'…the government is doing all the best they can do at the moment, as my colleague said that people just don't care about this virus. … I think they have now realised that it is really spreading very fast, and they are now taking all possible steps to stop this virus very quickly…'* [FG14 August 2020, Indian]

**Widespread testing:** *'Number two, and most importantly why isn't each and every house, we all go to a GP yeah, well why isn't, why didn't the Government ask every single person to go and have a COVID-19 test treated at your GP and not doing for certain people. …, from child to older people, you know every one, they could've of said, look go and have a test at your GP just like the way you have an appointment for any illness…'* [FG15 August 2020, male Bangladeshi].

**Belief that messaging is confusing and inconsistent:**

**Confusion: '***I'm a chairman of a patient's group and I get a lot of phone calls. What shall I do, etc, etc. So a lot of people are confused and there is no clarity…'* [FG18 August 2020, male, Indian].

*'Later on Dominic Cummins so clearly broke the rules, but the fact that he didn't publicly say that what he did was wrong it kind of meant that everyone was open to interpret it how they wanted to, so that for me was when it did get really unclear, because I was like, well maybe I can go for a drive, you know.'* [Mixed ethnicity FG2 June 2020, female].

**Prime ministers messaging** '*Especially the Prime Minister's message, I was totally, utterly, I was confused and so, it was really hilarious, sorry. I can't remember when was it but he was, he wasn't, he didn't know what he was saying so…It was really confusing. You can go to work, don't go to work, stay at home, I don't know. It was funny and I just laughed.'* [Mixed ethnicity FG1, June 2020, female].

'F*rustration is the best word that describes the way it was received by us. In terms of the messages, they were always very, unmatched to the situation. They were always upbeat and so positive, like we're literally doing so great, when things are quite critical. Also, it was very frustrating to watch. The Prime Minister kept stating like he's backing up everything with science, but I felt like he was very selective in choosing the messages that he was getting across.'* [FG11 July 2020, European female]

***A lack of consistency.*** *'You know like I was in, the eat out whatever it's called, help out eat out thing. I was in this pub and it was absolutely packed, no masks whatever, and then I walked to Sainsbury's and it's masks, it's wide, you know wide aisles and there's like a, how many people can go in, it just doesn't add up, you know like, it should be an equivocal policy that's going to help us reduce the numbers and help us get to a better state and it's not. It seems like there's a lot of kind of reactive people pleasing measures'* [FG22 September 2020, white British].

**Face masks:** *'One of my biggest concerns was about the whole face mask. Because I thought right at the beginning there was this whole argument, the science doesn't support us wearing face masks, and then just only recently that they've been telling us to go out and start wearing face masks. And then it makes me wonder, is it really because they're trying to get the people out and get the economy restarting. Is it really going to protect us or protect others? It's just, yes, I don't know, I feel like there's a bit of a trust issue.*' [FG10 July 2020, female, Bangladeshi].

**PPE:** *'in my industry, the biggest issue we had was PPE. At one point it was we don't need face masks and then all of a sudden, 12 weeks after the lockdown, everyone has to wear face masks.'* [Mixed ethnicity FG3 June 2020, male].

**Eat out to help out will spread the virus:** *'you know the Government started 50%, you know the discount if anyone eats inside the restaurant. there is no social distancing, it's over crowded, it's masks everywhere, where ever you go, you can see outside of the restaurants is long queue, people are waiting to go inside. So, they're going to be spreading the virus it's definitely, this is worse situation now.'* [FG15 August 2020, male Bangladeshi].

**Annoyance at government advisors breaking the rules:** *'even when the whole Dominic Cummings situation happened as well and Boris pretty much defended it and said you can do certain things, it's open to interpretation, I think it was at that, I guess that weekend, anything after that kind of just made me think, well we've literally been following your guidelines to a T and you've excused this behaviour, it was a bit of a kick in the teeth and it was, it was quite, it was really annoying…'* [Mixed ethnicity FG2 June 2020, female].

**Attitudes towards Public Health England:**

**PHE initially handled the pandemic like influenza:***'…It's been very clear to me from the beginning that Public Health England has failed to provide timely information (call drops) and accurate information, early enough, and in a sense whether this, whether there's been political pressure at the top to downplay things (call drops) again to treat this as Influenza, which isn't Influenza, as we all know. Yeah, so speaking among colleagues, a lot of scientists, a lot of microbiologists I know, and particularly those that are from an Asian background who come from Singapore or Hong Kong, who have experienced SARS in the past…'* [Mixed ethnicity FG3 June 2020, male.]

***Public Health England being used as a scapegoat by government:*** *Public Health England is doing the best they can, but politicians being politicians they scapegoat everybody but themselves. Everybody knows that, right? That's why people don't trust the government as opposed to the institutions, right? They can relabel Public Health England to whatever, to a Wizard of Oz nursery or whatever, right, it doesn't detract from the fact whether it's Mr Cummings, Mr Johnson, yeah, it's the same failings with any higher tier management…'* [FG17 August 2020, Chinese].

**Attitudes towards local government:**

**One positive attitude towards council***: '…for the first ten weeks I used to get called from the, either it's the health centre or the council, whatever it is, they used to ring me every week to find out if I was OK or if I needed any help, need any shopping or anything…'* [Male, FG9 July 2020, Indian].

**Lack of action to address COVID locally by council:***'that's why I'm a little bit critical about the government and the local council. The local council is for the local people and they're not putting their effort in to help the town* [FG18 August 2020, male, Indian].

*'… We were doing those kind of things, we had to do at our own back, but whereas council were busy funding little organisations that weren't doing anything, didn't really achieve much, that was annoying..'* [FG19 September 2020, South East Asian].

*' I think it's important that politicians, whether it's locally, nationally based, they look beyond the politics, so they're trying to look after, or trying to do things that are to help communities. Not further their own political agendas, or kind of agendas of individuals.'* [FG19 September 2020, South East Asian].

**Travellers views of councils:**' *they pass the buck and then nothing gets sorted.'* [FG24 October 2020, male, Traveller].

*'I mean I remember the council at one point turned round and said, oh, right we're going to open one toilet for you, we're going to open the toilet for you in *. And I remember going with somebody from FFT to see if they'd opened it and they'd opened it but they'd turned the tap off. So you could go to the loo but couldn't wash your hands and it's just, it was so dehumanising. It's just so dehumanising and I remember them saying, OK, well we've opened a tap up for you in the park, which is about the other side of *, six miles away from the functioning toilet. So we went over there and literally the only tap is just coming out the side of a wall by some bushes and it's just surrounded by rat traps and you could see rats scurrying around everywhere and there's rubbish everywhere and I just thought, do you know what, this is the most dehumanising treatment from such wealthy, progressive, liberal city.'* [FG24 October 2020, Traveller].

**ALTRUISM**

**Positive attitudes towards the NHS:**

*'from like watching on the media, they* [the NHS] *seem to have done an amazing job really and yeah, I think it's just that sort of like stoic kind of attitude that maybe the British have of like pulling together and yeah, like looking after people when times get tough and yeah, they, I think they've been amazing really, with what they've had to deal with.'* [FG22 September 2020, white British].

*'… I think the NHS have done perfectly. It's done all it can do you know, from a sort of, just an example. So, I got discharged from the hospital about three months' ago and I'm still having people ringing me up, getting me in for appointments to check up on me. I've had clinical psychologists checking me over. I've had multiple sisters calling* me to check up on me and if I'm honest, apart from a couple of xrays that have gone awry you know I can't, I can't fault the NHS, they've done all they can. If the Government's failing to sort of organise the healthy people, it's only their job to look after the sick people,

they can't do anymore and I really can't fault the NHS for what they've done…' [FG22 September 2020, male, white British].

**Negative attitude towards NHS:** '…I've never gone outside and clapped for the NHS because I don't particularly think my experience of them has been particularly good from the start of this going on with dealing with my mother…' [Mixed ethnicity FG3 June 2020, male].

**Contribution of MEGs:** *'And people from the communities were proud that their members were on the frontline.'* [FG19 September 2020, South East Asian].

**Community worked together** *'There was like all over it was so hard thing and stuff, but it just brought out the good in people and how they could work as a community and, you don't know how much someone could do, or that little thing, how can it make such different in your day.'* [FG13 August 2020, female, Arab].

**Families:** *'I don't know about other nationalities but from an Asian background, I think, from my household, we just took to the law that was provided and so we didn't really have to change our way. Yes, we might have to adjust and change a couple of things in terms of Ramadan, Eid and family get togethers but it's one of those things that, when a situation like this arises, that's what you have to do. If you're being told you have to do this then, so be it, you just have to go with it'* [Mixed ethnicity FG5 June 2020, male].

'*I use this as a, I'm going on a diet. I use this to fine tune my eating habits, my exercise habits, and so has my family. They've, rather than sit there and mope about it, we've gone to the positive side and tried to improve our lifestyle.'* [FG18 August 2020, male, Indian].

**TANGIBLE ACTIONS REQUESTED BY PARTICIPANTS**

**POLICIES**

**Policies that increase social equity** *'…I think it's a long term problem, I think until you've got equity and equality in terms of class and racism, I think it's, you're always going to have that health inequality in terms of poverty and just vulnerable people in society, so I think that is a long term problem that you need to dismantle these things in our society…'* [FG6 June 2020, Female, Pakistani].

*'I think there needs to be a massive overhaul of what is done to support people who are from the lower socio economic backgrounds because they are going to be those black and Asian kids, they are going to be those, the black and Asian mums and the dads who are probably going to be unemployed and now the children that are possibly, it's that perpetual cycle and I think that, it's what we do now that's important. It needs to be joined up thinking, it can't be ten years later or five years later'* [Mixed ethnicity FG5 June 2020, female].

**Healthcare policies to increase equity across ethnic groups:**

*'…The NHS clearly needs to change in how it functions and how it deals with things and it needs to be much more specific with how it deals with different races. Black people are different from Asian people, or different from Chinese people, and this BAME thing doesn't work. It doesn't work in, forget the context of this, it doesn't work generally and this just exacerbates the general societal problems…'* [Mixed ethnicity FG3 June 2020, male].

**Needs for black women during childbirth/ COVID-19 restrictions**: *' I always kind of knew about the stats that like, I think it's, black women are five times more likely to die in childbirth than white women, so I'm quite aware of that, I think you need a partner there to advocate for you and support you. So I do really feel sorry for women that were having to consider, yeah, giving birth without their partner.'* [Mixed ethnicity FG2 June 2020, mother who had baby just before lockdown].

**Clear guidance**:

'…a lot of people are confused and there is no clarity. You could have lots of reports, lots of things, but if they're not going to tackle the main issues (inaudible) …So we need some clear guidance, [FG18 August 2020, male, Indian].

*…But I need some more clarity from the government and the government needs to do something about the BAME community.'* [FG18 August 2020, male, Indian].

'T*he key workers, who are the doctors and nurses and a lot of BAME people fall into that category as well. So that's where PPE is very important …'* [Mixed ethnicity FG5 June 2020, female].

**Transparency:** *'…transparency would be, is really good moving forward, because it just means that people can make more informed decisions.'* [Female, FG8 July 2020, black ethnicities]
'*accountability with things that went wrong, so people to be held accountable for doing them.'* [*Mixed ethnicity FG2 June 2020*].
*'I think for me it's the underlying reason why our community is being affected, so we can then put strategies or be aware what's happening that's causing that, so to be able to deal with it*.' [*Mixed ethnicity FG2 June 2020*].

**One set of guidance does not fit all:**
*'For the government to communicate in different languages. So, a leaflet, for example, or something pushed through the door. Just to have the guidance in different languages would be really helpful.'* [FG12 August 2020 Chinese female].
*'They need to get the information out in a way that people can digest and, you know, in a much more targeted way. And I just think, from my point of view, the gap between clearly there was a problem with non-white people and that information coming out. It's only now that the information is there, one can make an informed decision and until then, there's a huge gap in that information coming out with recommendations that need to be done. So, to some extent it's about getting information out that's relevant.* [Mixed ethnicity FG3 June 2020, male].
*'This just highlights the differences, basically, how, in the, on one end of the spectrum you may have more BAME people in low socio economic status groups but on the other end you'll have people who are very well educated, who are the key workers, who are the doctors and nurses and a lot of BAME people fall into that category as well*.' [Mixed ethnicity FG5 June 2020, female].
***Approved of guidance for a Jewish festival.*** *'I think somebody has actually given it some thought as to how we, not how we operate but you know, how we function as a religion, and I find that quite comforting.…'* [FG21 September 2020, male, Jewish].
**Lack of need for specific religious guidance:**
*'…I don't think there's any specific support from the government just because I'm Jewish. As far as I'm concerned, I don't want to be singled out for being Jewish, because otherwise they're going to have to single out every other religion as well and see what they can do. And there's really nothing that I think they can do, that I know of, or that I can think of specifically because I'm Jewish. Why would I want to be different? Why would I want there to be an extra allowance for people going to the synagogue, maybe they can have 20 or 30 people going and then they can't in the Church of England or something like that…'* [FG21 September 2020, male, Jewish].
*Yes, I'm not religious at all, so for me, I don't think there's anything that would single me out in my religion or anything, I'd say that I would probably just do the same as every religion and, I, there's nothing that would say really other than that I could go to a meal at a family's house with more than six people, that is the only way it would probably affect my life in my religion. I'm not one of those people that goes to the synagogue every week, so it didn't have an effect on me previously anyway. I might have gone this weekend for the Jewish New Year once or twice, but that's about it, so yes…'* [FG21 September 2020, female, Jewish].
**Support of employment**:
'*when the Government launched the furlough scheme, our company was really happy and they wanted to keep everyone to make sure everyone is working, nobody is unemployed*. [FG15, male, Bangladeshi].
*'For me now I found it is employment,* if this is like over, *people like me even from not minority groups, they would have support to get back to work, or they would have an office to get help, a recruitment office or anything like that. Because it's going to be hard over such a long time, you're not being employed.'* [FG13 August 2020, female, Arab].

**INTERVENTIONS**
**Take actions once cause of risks is determined:**
*'If it does transpire that there are underlying reasons why they* [people] *are being more adversely affected that, that it is addressed. And even if it's not as straightforward as a vitamin D deficiency and it comes down to systematic problems* [then] *the government*

*have really committed to taking that and dealing with that.'* [*Mixed ethnicity FG2 June 2020, female*].

**Communication through different channels**

**Campaigns:** *And I think a few more campaigns just educating the public as well, because trying to disseminate information that is readable and digestible, because a lot of the time most people just get their information from the media, but they already have their own agendas and stuff. And I think trying to tackle that would really help people build confidence in the government and the organisations that are responsible for looking after their health*…' [FG8 July 2020, female, black].

**Radio: '**F*or example PNC's got their own radio, we could have played that message more effectively. Even though we didn't get the invitation, but we played our own message. And also to involve the community leaders from the Mosques and other places, that works better, in my opinion.'* [FG19 September 2020, South East Asian].

**IT support:***. 'they don't have access to technology, some don't know how to use technology, some of them they don't have money to have internet in their house. They don't have a proper equipment to [join] their support group now we are online.'* [FG23 September 2020, female black African].

**Work with local communities to increase understanding and optimise support:**

['*Short term I think, maybe outreach within council level would be really good for, that requires money which causes a problem, I don't know if that's something that can be done at this time, because I know that's an issue, but I think that would be really helpful for the community right now…'* [FG6 June 2020, female, Pakistani].

*'I would say all the learnings that we've had, in terms of you've got the grass roots people that you need to connect with, you've got the different cultures. You've got different living conditions, for some people who are living with extended families it might be much harder for them to self-isolate. So, it's understanding all the learnings that we have, and having a contingency plan ready and waiting if the second wave does come…'* [FG19 September 2020, South East Asian].

*'I think the BAME community does a lot to help out, not just the BAME community itself, and I think that would actually unite people more. I'm aware that but I mean I think Muslim charities and the Muslim community especially have been. '* [FG9 July 2020, female, Indian].

**Environmental restructuring:**

**Improve living standards for multigenerational households**: *'But the real question is, how can we avoid the communities of getting infected more. If I visit them in houses, I've seen flats where there are two different families with two or three kids each sharing three rooms. I don't see any sides ready to do what it really takes. The answer is systematic, … in order to actually do something really, really productive for them.'* [FG11 July 2020, Hispanic male]

**Travellers needed provision of basic services:**

'*I think it should be automatic that every authority throughout England, or Great Britain, should go out and provide a water bowser toilet, access to gas and wood…'* [FG24 October 2020, male, Traveller].

**Education for Healthcare workers around managing critical illness in different ethnic groups**

*'There is a big difference in the way healthcare workers like doctors and nurses and stuff look at patients who are BAME. I think especially because we're not really taught the differences from how a white person would present, as opposed to how someone of colour would. We don't look the same we wouldn't present the same so a lot of the time we find that people come in at later stages [*of illness] *because it's not as clear as it would be on a white person. And I think like we're not taught how things manifest differently, because of the colour of your skin, so I think that's why a lot of the time people are missed. And I think that's kind of a big thing with COVID as well, … we're not taught the differences so we have to go out of our way to see the differences, so I think that* [education around differences in presentation with illness] *would be a really good thing that Public Health could make sure happens…'* [Mixed ethnicity FG2 June 2020, female].

**Interventions to increase adherence to rules:**

'*policing the situation hasn't been right from the beginning, they could have been more stricter in everywhere, stricter in the inner city area where people are gathering together and they're behaving like there's nothing happening, and that's where the problem is occurring…'* [FG14 August 2020, male, Indian].

*'the government should have learnt from other countries and what they've done, bringing in curfews, very strict curfews and processes and systems. And I know that we have an open society here, but sometimes the strictness does need to, dare I say follow some of the Asian thinking…'* [FG14 August 2020, male, Indian].

*'I think the government should have been tougher on, I know they, it's really difficult to deliver all the penalties and things like that and the police just had to, they're on a hiding to nowhere really…we need somebody in charge, people are going to be like oh, there needs to be that respect there, which I think is just a wider issue that we've probably lost as a country. When I see a policeman walking down the street, I never think about throwing a bottle at him or anything like that like the, some people do today, I'm always, I'm still almost like really nervous because I think, oh my gosh am I doing something wrong. We need, you need to have figures in place which people are, they're almost scared of them, they actually believe that something will happen to them if they don't abide by the rules…'* [FG20 September 2020, female, white British].
